# *Chlamydia trachomatis* and *Neisseria gonorrhoeae* in newborns with and without neonatal conjunctivitis: cross-sectional study in Papua New Guinea

**DOI:** 10.64898/2026.07.09.26357364

**Authors:** Nicola Low, Alice Mengi, Lisa M Vallely, Claire Descombes, Lydia Braunack-Mayer, Mitchell Starr, Handan Wand, Ben D Spycher, Steven G Badman, Moses Laman, William S Pomat, Andrew J Vallely, Michaela A Riddell, the WANTAIM trial investigators

## Abstract

**Objectives:** *Neisseria gonorrhoeae* and *Chlamydia trachomatis* can be transmitted from an infected woman to her baby during vaginal birth to cause neonatal conjunctivitis (ophthalmia neonatorum). Little is known about how often these pathogens cause infection in babies without conjunctivitis. The objectives of this study were to investigate the frequency of *C. trachomatis* and *N. gonorrhoeae* in newborn babies with and without conjunctivitis and to estimate the prevalence of these infections and the proportion that was asymptomatic.

**Methods:** We conducted a cross-sectional study, nested in a randomised controlled trial in Papua New Guinea. At a visit scheduled 1–2-weeks after birth (visit 1), we took conjunctival swabs from babies with clinical signs of conjunctivitis and from a sample of those with no clinical signs. Babies with conjunctivitis received antibiotic treatment. Swabs were tested for *N. gonorrhoeae* and *C. trachomatis* by PCR after the end of the study. We estimated the weighted percentage (with 95% confidence intervals, CI) of babies with either organism detected and the asymptomatic proportion.

**Results:** From 21^st^ October, 2017 to 2^nd^ February, 2022, 1699 of 2055 newborn babies attended visit 1. Among 310 babies with clinical data and a result from any swab, 97 had signs of conjunctivitis and 213 did not. Where results were available, *C. trachomatis* was detected in 12/97 (12.4%) of babies with conjunctivitis and 7/211 (3.3%) without. *N. gonorrhoeae* was detected in 1/97 (1.0%) of babies with conjunctivitis and 6/210 (2.9%) without. The weighted estimate of *C. trachomatis* or *N. gonorrhoeae* prevalence was 6.6% (3.8-9.9%), of which an estimated 87.0% (74.8%-93.8%) was asymptomatic.

**Conclusions:** *C. trachomatis* and *N. gonorrhoeae* were detected in the absence of clinical signs of conjunctivitis in about 1 in 15 babies. This finding has implications for estimation of mother-to-child transmission and evaluation of prophylaxis and treatment interventions.

**Key messages:** *What is already known on this topic:*

▪ *Neisseria gonorrhoeae* and *Chlamydia trachomatis* cause neonatal conjunctivitis (ophthalmia neonatorum), which can lead to permanent vision loss if untreated;
▪ Little is known about the occurrence and frequency of *N. gonorrhoeae* and *C. trachomatis* in newborn babies without conjunctivitis.

*What this study adds:*

▪ This study included babies with clinical conjunctivitis and a sample of babies without clinical signs of conjunctivitis and tested swabs using PCR at a visit scheduled 1-2 weeks after birth;
▪ The overall prevalence of *C. trachomatis* or *N. gonorrhoeae* was estimated to be 6.6% (95% CI 3.8-9.9%), of which an estimated 87.0% (74.8%-93.8%) occurred in babies without clinical signs of conjunctivitis.

*How this study might affect research, practice or policy:*

▪ The clinical relevance of asymptomatic *C. trachomatis* or *N. gonorrhoeae* detection remains unclear; all samples from babies with asymptomatic detection tested negative at a visit scheduled 4-6 weeks after birth;
▪ The results are important for research about mother-to-child transmission of *C. trachomatis* and *N. gonorrhoeae* and evaluation of prophylaxis and treatment interventions conducted using molecular diagnostics.

## Introduction

The sexually transmitted bacteria *Chlamydia trachomatis* and *Neisseria gonorrhoeae* can be transferred from an infected pregnant woman to the eyes of a newborn during vaginal birth. Both can cause neonatal conjunctivitis (ophthalmia neonatorum),^1-3^ which can result in permanent vision loss if untreated.^1^ Antibiotic eye ointment at birth is recommended to prevent chlamydial and gonococcal neonatal conjunctivitis.^1^

Much of our knowledge about gonococcal and chlamydial neonatal conjunctivitis comes from studies that used culture for diagnosis.^2 3^ The frequency of asymptomatic detection is not known because studies tend to include only babies diagnosed with conjunctivitis.^2^ Nucleic acid amplification tests are now the recommended assays for detection of *C. trachomatis* and *N. gonorrhoeae* in adults.^4^ Although not formally validated, they should detect these organisms in conjunctival specimens. The objectives of this study were to: 1) determine the frequency of *C. trachomatis* and *N. gonorrhoeae* in newborns with and without conjunctivitis, and 2) estimate the overall prevalence of *C. trachomatis* and *N. gonorrhoeae* detected in the eyes of newborns and the fraction that is asymptomatic.

## Methods

We report this study according to the Standards of Reporting for Observational studies in Epidemiology (online checklist).

This study was nested in the Women and Newborns Trial of Antenatal Interventions and Management (WANTAIM) in Papua New Guinea (PNG), which evaluated antenatal screening for sexually transmitted infections (STIs) among women enrolled at 10 study clinics.

WANTAIM-neonatal was a sub-study conducted at five clinics, selected at random. The study received approval from the PNG Medical Research Advisory Committee (MRAC.16.24), PNG Institute of Medical Research (IRB.1608), University of New South Wales (HREC.16708), London School of Hygiene and Tropical Medicine (REC.12009) and the Canton of Bern (2021-01209). The protocol and main trial findings have been published.^5 6^

Research staff asked pregnant women who agreed to take part in WANTAIM if they would allow extended follow-up of their baby beyond the postnatal examination, which took place for all babies. If the woman gave informed consent, two additional visits were scheduled; visit 1, 1–2 weeks after birth and visit 2, 4–6 weeks after birth. At each study visit, trained staff collected questionnaire data, examined the baby and collected samples. Following national guidance and availability of supplies, all babies born or examined at a health facility soon after birth received prophylactic oxytetracycline or chloramphenicol eye ointment and all babies with conjunctivitis received treatment.

All babies attending study visit 1 were eligible for this study. This time window allowed for diagnosis of conjunctivitis, whilst reducing detection of bacterial nucleic acid from maternal secretions. Before the study started, we selected 15% of all identification numbers at random, which allowed examination of a sub-sample of babies without conjunctivitis, rather than taking swabs from all asymptomatic babies. At visit 1, study staff were instructed to take swabs from any baby with a selected identification number and from babies with any of four signs of conjunctivitis in either eye; purulent discharge, swelling, redness, or watery/teary eyes. They used paediatric flocked swabs (Copan, Brescia, Italy) to take two samples from the lower conjunctiva of each eye. They placed one set (left and right eye) into cobas^®^ PCR Media (Roche Diagnostics, Rotkreuz, Switzerland) and one set into GeneXpert^®^ Swab Transport Reagent (Cepheid, Sunnyvale, CA, United States of America). All swabs were stored in a cooler at the clinic and transported daily to the laboratory. Swabs for Xpert CT/NG were stored at 4°C and tested the following day in PNG. Cobas samples were stored at -80°C until transport to St. Vincent’s Hospital, Sydney, Australia; they were tested on the cobas 6800 platform after the end of the study. Tests were performed according to the manufacturer’s instructions. Swabs were subsequently used in studies of diagnostic accuracy and mother-to-child-transmission, which will be reported separately.

We analysed data from all babies who attended visit 1, had a clinical eye examination and valid PCR result. We defined conjunctivitis as: purulent discharge in either eye or the presence of two or more of swelling, redness, watery/teary eye. We defined detection of *C. trachomatis* or *N. gonorrhoeae* as a positive result using either cobas CT/NG or Xpert CT/NG. If test results were discordant, we used the cobas PCR result. We first described detection of *C. trachomatis* or *N. gonorrhoeae* among babies with conjunctivitis and among the sub-sample without conjunctivitis as percentages with 95% confidence intervals (CI). To estimate a) the overall prevalence and b) the proportion of asymptomatic *C. trachomatis* or *N. gonorrhoeae* detected, we used the sampling fraction of babies without conjunctivitis to calculate weighted proportions, reflecting what would have happened if eye swabs had been taken from all babies. We calculated 95% CIs from 2,000 bootstrapped replicates (full statistical analysis plan, supplemental text S1).

## Results

The 2055 babies enrolled were born from October 21, 2017, to February 2, 2022, and 1979/2050 (96.5%, 5 missing data) received antibiotic eye ointment. Of all enrolled babies, 1699/2055 (82.7%) attended visit 1 at a median of 11 days (interquartile range 8, 13) after birth. Among all babies attending visit 1, 97/1699 (5.7%) had a diagnosis of conjunctivitis (supplemental figure S1). No adverse events were reported.

Of babies with conjunctivitis, 12/97 (12.4%) had *C. trachomatis* detected and 1/97 (1.0%) had *N. gonorrhoeae* (Table 1). Among the sample of babies without conjunctivitis and a test result available, 7/211 (3.3%) had *C. trachomatis* detected and 6/210 (2.9%) had *N. gonorrhoeae*.

**Table 1:** Positivity of *C. trachomatis* and *N. gonorrhoeae* in lower conjunctival swabs, by PCR at study visit 1, among all babies with conjunctivitis and a sample of those without.

| Pathogen | Conjunctivitis | # detected / # tested | Positive, % (95% confidence interval) |
|---|---|---|---|
| *C. trachomatis* | Present | 12/97 | 12.4 (6.6-20.6) |
| .. | Absent | 7/211 | 3.3 (1.3-6.7) |
| *N. gonorrhoeae* | Present | 1/97 | 1.0 (0.0-5.6) |
| .. | Absent | 6/210 | 2.9 (1.1-6.1) |

Based on the numbers of positive tests among all babies with conjunctivitis (13/97) and among the sub-sample of babies without conjunctivitis (13/211), the weighted estimate of *C. trachomatis* or *N. gonorrhoeae* prevalence was 6.6% (3.8-9.9%). The weighted estimate of the asymptomatic fraction of *C. trachomatis* or *N. gonorrhoeae* was 87.0% (74.8 %-93.8%).

Among 25 babies at visit 1, 18 had *C. trachomatis* and seven had *N. gonorrhoeae* (supplemental figure S2). Of 12 babies with chlamydial conjunctivitis, 10 received chloramphenicol ointment for treatment; reasons for non-treatment of two babies were not documented. One baby with gonococcal conjunctivitis received oral erythromycin. Twenty-one babies attended visit 2. Two babies with chlamydial conjunctivitis had *C. trachomatis* detected again. Both had received chloramphenicol eye ointment; one still had conjunctivitis and received oral erythromycin, the other no longer had conjunctivitis and received no further treatment.

## Discussion

Among 2055 newborn babies, almost all received ocular antibiotic prophylaxis. At a visit scheduled 1-2 weeks after birth, 13/97 (13.4%) with conjunctivitis and 13/211 (6.2%) of a sub-sample without conjunctivitis had *C. trachomatis* or *N. gonorrhoeae* detected. We estimated that 6.6% (95% CI 3.8-9.9%) of all babies would have had *C. trachomatis* or *N. gonorrhoeae* detected, of which 87.0% (95% CI 74.8-93.8%) was asymptomatic.

The main strengths of this study are the large overall sample size and inclusion of a sample of babies without clinical signs, which avoided the need to take swabs from all babies. Although some babies with eye swabs taken were not part of the pre-specified random sample, we could not determine any reason for systematic bias. A further limitation is the scheduling of a visit 1–2 weeks after birth. Positive results for *C. trachomatis* and *N. gonorrhoeae* could be false positives, including possible detection of nonviable nucleic acid,^4^ which would overestimate infection prevalence. There are also potential reasons for underestimation. First, ocular prophylaxis prevented some infections. Second, we might have excluded some cases of gonococcal conjunctivitis, given the short incubation period,^1 2^ if infection resolved or was treated before visit 1.

We found that most PCR-detected *C. trachomatis* and *N. gonorrhoeae* occurred in the absence of clinical conjunctivitis. We found one small study of asymptomatic neonatal *C. trachomatis*, detected by culture in Nairobi, Kenya.^3^ Two of 49 babies (4.1%) had *C. trachomatis* detected at a visit seven days after birth, and both had conjunctivitis. Seven of 11 (64%) babies with *C. trachomatis* detected up to six months of age did not have conjunctivitis and no ocular complications occurred. In Botswana, researchers tested babies of 26 mothers with *C. trachomatis* and *N. gonorrhoeae* using GeneXpert.^7^ Seven had *C. trachomatis* or *N. gonorrhoeae* in eye swabs but the asymptomatic fraction could not be calculated.

The clinical implications of asymptomatic neonatal ocular *C. trachomatis* and *N. gonorrhoeae* detection remain unclear. In our study, all detected *C. trachomatis* or *N. gonorrhoeae* in babies without conjunctivitis had resolved by visit 2, among 21/25 who returned. In further research studies, the proportion of positive results caused by nonviable nucleic acid could be investigated using viability PCR assays.^8^ Our findings will contribute to contemporary estimates of mother-to-child transmission of *C. trachomatis* and *N. gonorrhoeae* among babies with and without clinical signs of infection.^6^ There is a need to review antibiotics used for ocular prophylaxis in PNG. Oxytetracycline is active against *C. trachomatis* but chloramphenicol may not be.^9^ Tetracycline resistance to *N. gonorrhoeae* might have emerged since the last assessment in 2010^10^ and susceptibility to chloramphenicol is unknown. In summary, *C. trachomatis* and *N. gonorrhoeae* were not the most common causes of neonatal conjunctivitis among babies who received ocular prophylaxis, but most PCR-detected *C. trachomatis* and *N. gonorrhoeae* occurred in the absence of clinical conjunctivitis.

## Supporting information

checklist

## Data Availability

All data produced in the present study are available upon reasonable request to the authors

## Acknowledgements

We thank the women who participated in the WANTAIM Trial, the families and communities who supported them and the staff who worked on the WANTAIM Trial (supplemental text S2). We thank Prof. Philip H Cunningham for his supervision of the laboratory testing and Mr. Andrew Kelly for his help with confirmation of results at St. Vincent’s Hospital Sydney, and we acknowledge the contributions of the following members of the WANTAIM trial investigator team: Prof. Caroline Homer, Prof. Rebecca Guy, Prof. John Kaldor, Assoc. Prof. Stanley Luchters, Prof. Glen Mola, Dr. Grace Kariwiga, Prof. Virginia Wiseman, Dr. Chris Morgan, Prof. Stephen Rogerson, Prof. Sepehr Tabrizi, Assoc. Prof. David Whiley, Prof.

Suzanne Garland, Prof. Rosanna Peeling, Prof. Peter Siba, Dr. John Bolnga, Dr. Leanne Robinson, Dr. Jacob Morewaya, Dr. Delly Babona, Dr. Neha Batura, Assoc. Prof. Angela Kelly-Hanku, Dr. Pamela Toliman, Mr. Wilfred Peter, Dr. Liz Peach, Ms. Lucy Au, Ms. Irene Pukai Gani.

## Author contributions

Conceptualization: NL, ML, WSP, AJV; Data Curation: CD, LB-M, HW, BDS, MAR; Formal Analysis: CD, LB-M; Funding Acquisition: NL, LMV, ML, WSP, AJV; Investigation: NL, AM, LMV, MS, MAR; Methodology: NL, LMV, CD, LB-M, AJV, MAR; Project

Administration: AM, LMV, SGB, AJV, MAR; Resources: NL, LMV, AJV, MAR; Software: CD, LB-M, HW, BDS, MAR; Supervision: NL, LMV, SGB, ML, AJV, MAR; Validation: NL,

LMV, CD, LB-M, HW, BDS, MAR; Visualization: CD, LB-M; Writing – original draft: NL,

LMV, CD, LB-M, MAR; Writing – review & editing: NL, AM, LMV, CD, LB-M, MS, SGB, MAR.

## Data availability statement

The data underlying this article will be shared on reasonable request to the corresponding author.

