## supplemental for "*Chlamydia trachomatis* and *Neisseria gonorrhoeae* in newborns with and without neonatal conjunctivitis: cross-sectional study in Papua New Guinea"

#### Contents

| Item | | Page |
|---|---|---|
| Figure S1 | Flow chart of babies enrolled in WANTAIM-neonatal with attendance at visit 1, according to diagnosis of conjunctivitis and availability of eye swab results | 2 |
| Figure S2 | Outcome of newborns with *C. trachomatis* or *N. gonorrhoeae* detected at visit1, when followed up at a visit scheduled 4-6 weeks after birth (visit 2) | 3 |
| Text S1 | WANTAIM-neonatal. Statistical analysis plan, version 1.3 | 4-17 |
| Text S2 | WANTAIM Trial study team members and project staff | 18-22 |

#### Supplementary material

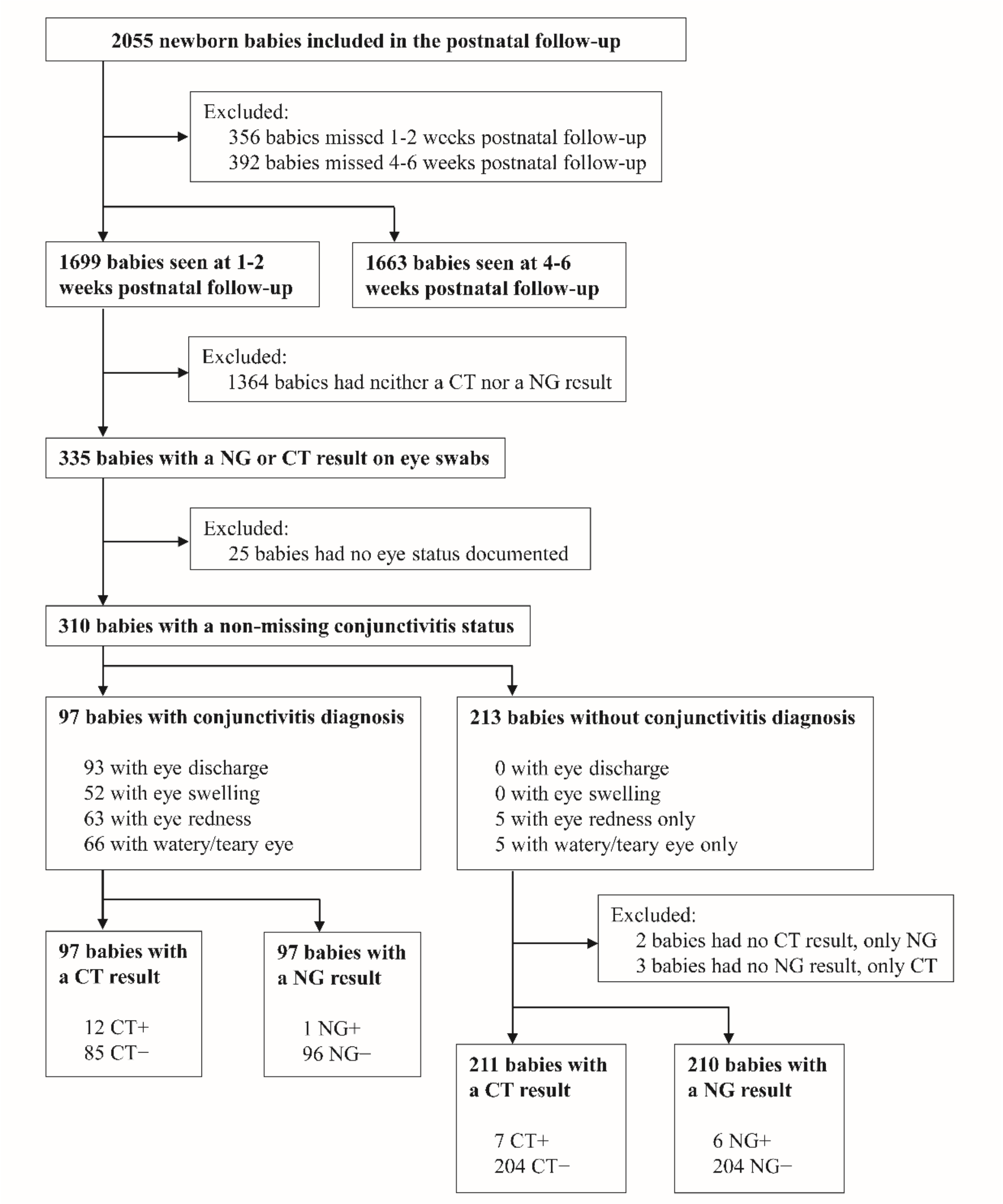

Figure S1: Flow chart of babies enrolled in WANTAIM-neonatal with attendance at visit 1, according to diagnosis of conjunctivitis and availability of eye swab results.

CT, *C. trachomatis*; NG, *N. gonorrhoeae.*

#### Supplementary material

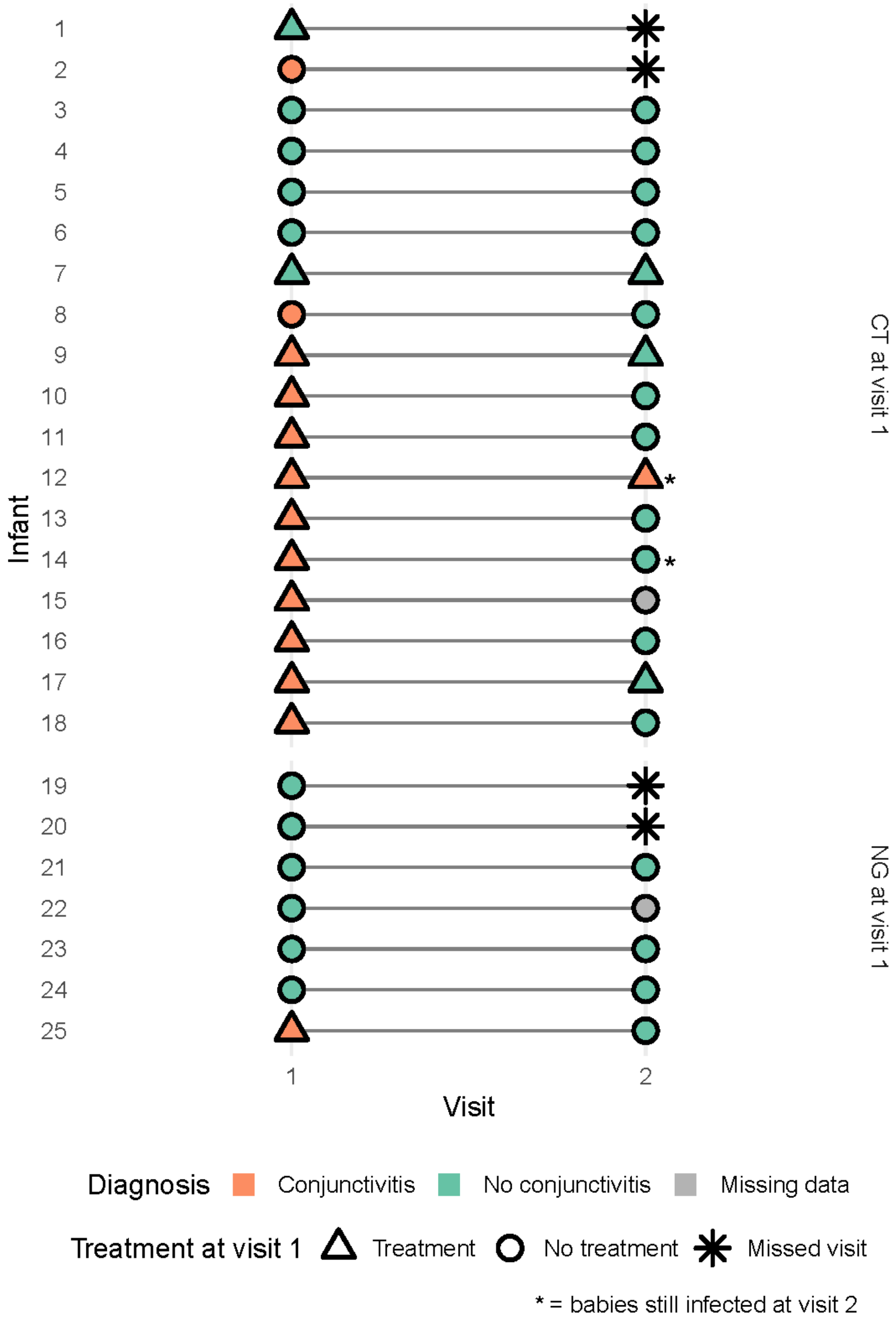

Figure S2: Outcome of newborns with *C. trachomatis* or *N. gonorrhoeae* detected at visit1, when followed up at visit 2, scheduled 4-6 weeks after birth

CT, *C. trachomatis*; NG, *N. gonorrhoeae.*

### WANTAIM-neonatal

#### Statistical analysis plan

Edited and expanded extract from the statistical analysis plan (SAP v9.0, 29 Oct 2022) for the WANTAIM clinical trial

### 1. Administrative information

| | |
|---|---|
| **Project title** | WANTAIM neonatal |
| **Trial registration number** | ISRCTN Registry, ISRCTN37134032 |
| **SAP version** | V1.0 (02.02.2026) |
| **Protocol version** | Vallely AJ, et al. Point-of-care testing and treatment of sexually transmitted infections to improve birth outcomes in high-burden, low-income settings: Study protocol for a cluster randomized crossover trial (the WANTAIM Trial, Papua New Guinea). Wellcome Open Res. 2019. doi: 10.12688/wellcomeopenres.15173.2 |

#### Contributors

| Name | Affiliation | Role in SAP writing |
|---|---|---|
| Prof Ben Spycher | ISPM, University of Bern | Author |
| A/Prof Handan Wand | Kirby Institute, University of New South Wales | Author |
| Dr Lydia Braunack-Mayer | ISPM, University of Bern | Author |
| Claire Descombes | ISPM, University of Bern | Author |
| Prof Nicola Low | ISPM, University of Bern | Principal Investigator |

#### Revision history

| Version | Section | Date | Summary of changes |
|---|---|---|---|
| 1.0 | | 02.02.2026 | First version |
| 1.1 | Objectives; Study methods; Statistical analyses | 27.05.2026 | Technical editing and proofing, clarification of definitions of clinical conjunctivitis and eye infection. |
| | Flow chart of mother-baby pairs and descriptive characteristics | | Clarification that flow charts will be created for both the full study population and control population. |
| 1.2 | Horvitz-Thompson estimator | 12.06.2026 | The Horvitz-Thompson estimator of prevalence is corrected due to incorrect weighting, and a similar estimator was constructed to estimate the proportion of asymptomatic babies among babies with a positive test result. |
| | Study population for prevalence estimation | 15.06.2026 | The study population used to estimate the prevalence of neonatal infection at visit 1 is corrected from 'control population' to 'full study population', since there is no reason not to use |

| | | | |
|---|---|---|---|
| | | | the full study population as long as it does not also include the mothers. |
| 1.3 | Estimation of transmission | 09.09.2026 | The computation of the 95% confidence interval (CI) for the transmission proportion did not account for the fact that some mothers had twins. This introduces a correlation that must be considered when calculating the CI. Clopper-Pearson confidence intervals have been replaced with cluster-robust 95% CIs (GLM + sandwich HC2). |

#### Objectives

The Women and Newborns Trial of Antenatal Interventions and Management (WANTAIM) clinical trial has three secondary outcome measures evaluated as part of the WANTAIM neonatal study. This document is the statistical analysis plan for: **Objective 8,** to measure the incidence of mother to child transmission of *C. trachomatis* (CT) or *N. gonorrhoeae* (NG) as indicated by positive newborn eye (CT or NG) or nasopharyngeal (CT) swabs by the 4-6 weeks postnatal visit; **Objective 9**, to measure the diagnostic test accuracy of the Xpert CT/NG assay compared with laboratory-based PCR.

#### Study methods

##### Trial design and setting

WANTAIM neonatal (previously referred to as neoSTI) was conducted in 5 of the 10 clusters in the main WANTAIM cluster-randomised controlled crossover trial in two provinces in PNG (Madang and East New Britain).[1,2] The clusters were selected at random before the start of the trial.

##### Eligibility and exclusion criteria

Women enrolled in the participating clusters were asked at enrolment for permission for their newborn baby to have an extended period of follow-up after the postnatal visit. We included all babies who attended the immediate postnatal visit (see section 3.3). We excluded liveborn babies who died before any of the scheduled postnatal visits.

##### Study visits and outcomes assessed

Participants in neoSTI had a postnatal visit (which all WANTAIM participants had, referred to as 'visit 0' or 'postnatal visit') and two additional follow-up visits, scheduled at 1-2 weeks after birth (referred to as 'visit 1' or '1-2 weeks visit') and 4-6 weeks after birth (referred to as 'visit 2' or '4-6 weeks visit') (Table 1).

**Table 1.** Samples collected, by visit, from babies born to women in control group, from standard operating procedures collection of eye swabs and for collection of nasopharyngeal swabs

| Outcome | Postnatal visit, visit 0 | 1-2 weeks visit, visit 1 | 4-6 weeks visit, visit 2 |
|---|---|---|---|
| **Eye infection** | | | |
| *Clinical examination* | L and R eye | L and R eye | L and R eye |
| Eligible | All enrolled | All enrolled | All enrolled |
| *Swabs taken* | L and R eye | L and R eye | L and R eye |
| Eligible | All enrolled | Signs of eye infection + 15% random sample, irrespective of signs | Signs of eye infection .. |
| Sample 1 | Swab each eye separately, 2 swabs in 1 tube | Swab each eye separately, 2 swabs in 1 tube | Swab each eye separately, 2 swabs in 1 tube |

| Outcome | Postnatal visit, visit 0 | 1-2 weeks visit, visit 1 | 4-6 weeks visit, visit 2 |
|---|---|---|---|
| Assay 1*<br>Sample 2<br>Assay 2† | Roche PCR (Sydney)<br>..<br>.. | Roche PCR (Sydney)<br>2 swabs in 1 tube<br>GeneXpert (PNG) | Roche PCR (Sydney)<br>2 swabs in 1 tube<br>GeneXpert (PNG) |
| **Pneumonia**<br>*Clinical examination*<br>Eligible<br>Sample 1<br><br>Assay 1*<br>Sample 2<br>Assay 2† | <br>Respiratory system<br>All newborns<br>..<br><br>..<br><br>.. | <br>Respiratory system<br>All newborns<br>Swab, left nostril<br>1 swab in 1 tube<br>Roche PCR (Sydney)<br>1 swab in 1 tube<br>GeneXpert (PNG) | <br>Respiratory system<br>All newborns<br>Swab, left nostril<br>1 swab in 1 tube<br>Roche PCR (Sydney)<br>1 swab in 1 tube<br>GeneXpert (PNG) |

PNG, Papua New Guinea; Sydney, St. Vincent's Hospital, Sydney, Australia

* Roche CT/NG PCR assay for *C. trachomatis* and *N. gonorrhoeae,* Roche cobas 6800 platform (Roche Diagnostics, Rotkreuz, Switzerland);

† Xpert CT/NG assay for *C. trachomatis* and *N. gonorrhoeae*, GeneXpert platform (Cepheid, Sunnyvale, California, United States of America).

#### Postnatal visit, visit 0

Visit 0 was scheduled within 72 hours after birth (Figure 1). All newborns had a clinical examination, and chloramphenicol or tetracycline eye ointment was applied, after taking swabs where possible.[1] All newborns had eye swabs taken from their left and right lower conjunctiva, placed in the same specimen tube (Table 1). Swabs were stored at minus 80 Celsius and tested by PCR (cobas CT/NG assay) at St. Vincent's Hospital, Sydney, Australia, after the end of the study. Swabs for testing on the GeneXpert platform were not taken at this visit.

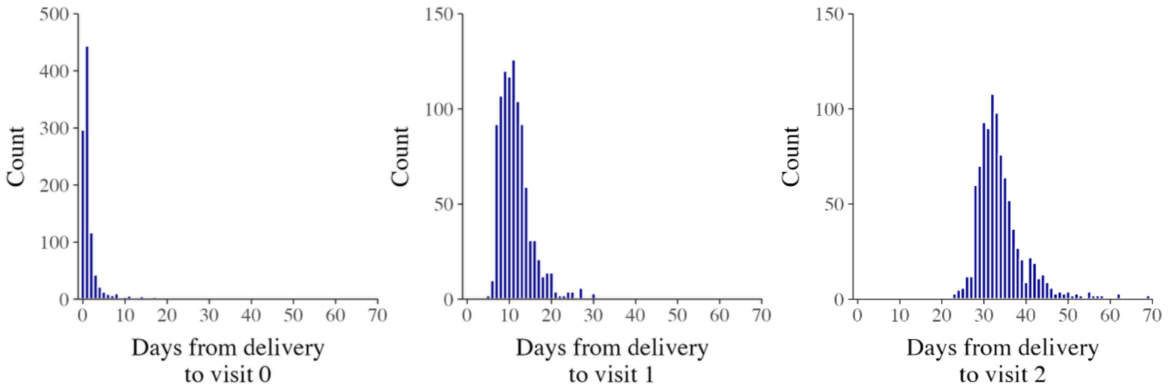

Figure 1: Days from delivery to each visit

#### Follow-up visit 1

Visit 1 was scheduled 1-2 weeks after birth. All newborns had a clinical examination and two nasal swabs taken from their left nostril (Table 1). Nasal swabs were tested in PNG by Xpert CT/NG assay (GeneXpert), and were stored and tested at St. Vincent's Hospital, Sydney, after the end of the study. If the newborn had clinical signs of conjunctivitis, eye swabs were taken from the left and right lower conjunctiva. One set of swabs was placed in the same specimen tube and tested by GeneXpert in PNG, with treatment if the test result was positive. One set of swabs was stored at minus 80 Celsius and tested at St. Vincent's Hospital, Sydney, Australia, after the end of the study.

Before the study started, 15% of identification numbers allocated to newborns were selected at random. At visit 1, any baby allocated to this subset was assigned to have eye swabs taken, even if they did not have clinical signs of conjunctivitis. Using the same procedure as for newborns with signs of conjunctivitis, eye swabs were taken from the left and right lower conjunctiva. One set was placed in the same specimen tube and tested by GeneXpert in PNG, with treatment if the test result was positive. One set was stored at minus 80 Celsius and tested at St. Vincent's Hospital, Sydney, Australia, after the end of the study.

##### Follow-up visit 2

Visit 2 was scheduled 4-6 weeks after birth. All newborns had a clinical examination and two nasal swabs were taken from their left nostril (Table 1). Nasal swabs were tested in PNG by GeneXpert, and were stored and tested at St. Vincent's Hospital, Sydney, after the end of the study. If the newborn had clinical signs of conjunctivitis, eye swabs were taken from their left and right lower conjunctiva. One set was placed in the same specimen tube and tested by GeneXpert in PNG, with treatment if the test result was positive. One set of swabs was stored at minus 80 Celsius and tested at St. Vincent's Hospital, Sydney, Australia, after the end of the study.

##### Eye infection

At any eligible visit, conjunctivitis is defined as the presence of discharge in either eye, with or without any other clinical sign, or any two or more of the following signs, in either eye: redness, swelling, teary/watery eye. Eye infection is defined as the detection of *C. trachomatis* or *N. gonorrhoeae*, or both, in an eye swab.

#### Study populations

We will consider three study populations for analysis:

- **Full study population:** mothers and newborns who attended at least one postnatal visit.
- **Control population:** mothers and newborns who attended at least one postnatal visit, and who were part of the control group of the WANTAIM parent study.
- **Random 15% population:** the 15% of newborns in the control group who were randomly selected to have eye swabs collected at visit 1 even if they did not have clinical signs of conjunctivitis.

#### Missing data

Analyses will be performed on complete cases only; babies with missing information for a given analysis will be excluded. No imputation will be performed. The number and proportion of missing values for each variable will be reported.

#### Statistical analyses

Unless otherwise specified, statistical analyses of mother-to-child-transmission will be done among mothers and their newborns in the **control population** because these mothers did not receive aetiological testing for STIs during pregnancy. They received syndromic management if they had symptoms of vaginal discharge syndrome, which would have resulted in treatment in a small proportion of STIs.[2]

Throughout all analyses, we will not adjust for the phase or cluster allocation of the WANTAIM randomised control trial. Our objectives are to estimate statistics within the study population, rather than to generate estimates that are representative of the wider PNG population. In addition, there is no plausible source of confounding by study phase, so adjustment for phase is not required.

##### Flow chart of mother-baby pairs and descriptive characteristics

For the **full study population** and for the **control** population, we will create a flow chart to show the numbers of mother-baby pairs at the start of the study, reasons for exclusion, the number analysed, and the number present for each visit.

Tables will show summary statistics of essential characteristics of mothers and of their babies. Categorical characteristics will be summarised as the number and percentage (n/N mother-baby pairs analysed). Continuous characteristics will be summarised with a median and interquartile range.

Essential characteristics for mothers include:

- Age
- Parity
- Marital status
- Highest educational level achieved
- Employment
- Smoking
- Alcohol consumption
- Betel nut consumption
- Body Mass Index ($kg/m^2$)
- Mid upper arm circumference (cm)
- Gestation age at enrolment (weeks)
- Haemoglobin (g/L)

- Malaria
- Syphilis test result
- HIV test result
- *C. trachomatis, N. gonorrhoeae* and *Trichomonas vaginalis* test results at 3rd trimester visitsEssential characteristics for newborns include:
- Receipt of eye ointment
- Multiple births
- Preterm birth (<37 weeks)
- Low birth weight (<2500g)

#### Newborn test positivity

We will calculate the number and percentage (n/N total newborns) of newborns with a positive *C. trachomatis* or *N. gonorrhoeae* result. These calculations will be done for results of eye and nasal samples, for each pathogen, for babies with and without clinical signs of conjunctivitis, and for babies with and without clinical signs of pneumonia.

We will also tabulate the number and percentage of newborns with a positive *C. trachomatis* or *N. gonorrhoeae* result among newborns in the **random 15% population** (n/N newborns in the **random 15% population**).

#### Cross-tabulation of concordance

##### Concordance between maternal and newborn results

The concordance between maternal and newborn results will be calculated. For the babies, a positive test result at any visit is counted as positive, and those with all negative results are defined as negative. Positive results are positive by Roche cobas (Sydney) or, if no result from Sydney, by GeneXpert

These calculations will be done for results of eye and nasal samples and for each pathogen, based on 2x2 tables (Table 4). Separate tables will be made for newborn eye swab results and nasal swab results for: *C. trachomatis*; *N. gonorrhoeae*; both *C. trachomatis* or *N. gonorrhoeae*. Separate tables will be made for each study visit and for cumulative result by the end of follow up, following the format in Table 2.

Table 2. Concordance between mother and baby test results, overall

| | Mother result* | | |
|---|---|---|---|
| Baby result* | Negative | Positive | Total |
| Negative | a | b | a+b |
| Positive | c | d | c+d |
| Total | a+c | b+d | N |

* Order of test results is the same as Stata output when 0=negative, 1=positive. Order of negative and positive might need to be changed for publication

##### Concordant results: mother positive, baby positive

The proportion with positive concordant results (d/b+d) will be calculated with Clopper-Pearson 95% confidence intervals (CI).

At the first visit after birth, we will use the term ‘positive result’ in the newborn because it is not known whether a positive result reflects transfer of organism from the vagina during delivery with transient colonisation, or infection. At the follow-up visits scheduled at 1-2 and 4-6 weeks, we will interpret a positive test result in an eye swab as infection. A positive nasal swab result for *C. trachomatis* may be colonisation or infection. The interpretation of a positive nasal swab result for *N. gonorrhoeae* is unclear.

##### Discordant results: mother negative, baby positive

The proportion with discordant results (c/a+c) will be calculated with Clopper-Pearson 95% confidence intervals.

Discordant results, with a negative test in the mother and positive test in the baby could represent a false negative test in the mother, a missed infection in a mother who acquired an infection between the third trimester visit and delivery, or a false positive result from the test in the baby’s sample. False positive results in the Roche cobas and GeneXpert assays are considered to be <1%.

These results will be examined individually and the individual results from Roche cobas and GeneXpert will be reported. The presence of clinical signs of conjunctivitis will also be examined for babies with a positive test for whom mothers had a negative test result.

#### Concordance between newborn test results and clinical outcomes

At the postnatal visit (visit 0), at least, there should be high concordance between a positive test result for *C. trachomatis* or *N. gonorrhoeae* and clinical conjunctivitis.

*Table 3. Concordance between newborn test results and clinical outcomes*

| | Clinical outcome* | | |
|---|---|---|---|
| Baby result* | Negative | Positive | Total |
| Negative | a | b | a+b |
| Positive | c | d | c+d |
| Total | a+c | b+d | N |

* Order of test results is the same as Stata output when 0=negative, 1=positive

The proportion with positive agreement results (d/b+d) will be calculated with Clopper-Pearson 95% confidence intervals (CI).

#### Mother-to-child transmission

The overall proportion of transmission, for each pathogen and each sample type, will be calculated in two ways:

- The proportion (with cluster-robust 95% CI (GLM + Sandwich HC2)) of babies with a concordant positive result (d) out of all mothers with a positive result in the third trimester (b+d) (Table 2). This is a minimum estimate.
- The proportion (with cluster-robust 95% CI (GLM + Sandwich HC2))) of all babies with any positive result (c+d) out of all mothers with a positive result in the third trimester or with a negative result where their baby was positive (b+c+d) (Table 2). This is a maximum estimate.

These approaches come with limitations. For eye swabs, not all babies were tested at the 1-2 week and 4-6 weeks visits. Positive results would only be able to be detected in babies with clinical signs (or in the **random 15% population** tested at 1-2 weeks). It would be assumed that any baby not tested was negative for both pathogens, which might underestimate both infection prevalence in babies and concordance.

For nasal swabs, no nasal swab was taken at the postnatal visit. This is not a serious problem, because transient positivity is probably not established infection. Positive results for both *C. trachomatis* and *N. gonorrhoeae* at either the 1-2 weeks or 4-6 weeks visits can be considered nasal carriage. For *C. trachomatis*, nasal carriage is presumed to put the baby at risk of pneumonia. The clinical significance of *N. gonorrhoeae* nasal carriage is unknown, but we can report it.

#### Prevalence of neonatal infection

##### Prevalence of neonatal infection at visit 1

All available test results from the **full study population** will be used to estimate the prevalence of asymptomatic *C. trachomatis* and *N. gonorrhoeae* at 1-2 weeks. This will include eye swabs taken both from babies with clinical signs of conjunctivitis and from babies from the **random 15% population**, without clinical signs of conjunctivitis (described in section 4). This proportion can be applied to all babies to obtain an estimate of overall prevalence at the 1-2 weeks visit, given the following assumptions:

- **Assay performance**: GeneXpert and Roche Cobas have high sensitivity and specificity on eye samples from babies. Few infections will be missed, and there will be few false positives.
- **Adherence to protocol:** babies without signs of conjunctivitis should only have been sampled if they were part of the random 15%. This calculation will use all babies who have complete information on both clinical outcomes of eye infection and test results. For babies without signs of conjunctivitis, this may include more than just infants recruited into the **random 15% population** (babies sampled without clinical signs of conjunctivitis) and so may not constitute a random sample.

For each pathogen, prevalence at visit 1 will be estimated using a Horvitz-Thompson estimator[3] to weight according to the sampling design:

$$\frac{1}{M}\left(w_{asymptomatic} \times c + w_{symptomatic} \times d\right)$$

For $w_{asymptomatic} = \frac{1}{15\%}$, $w_{symptomatic} = 1$ and $M = (a + c) \times w_{asymptomatic} + (b + d) \times w_{symptomatic}$.

For each pathogen, proportion of asymptomatic babies among babies positive to that pathogen at visit 1 will be estimated using a Horvitz-Thompson estimator to weight according to the sampling design:

$$\frac{1}{M}\left(w_{asymptomatic} \times c\right)$$

For $w_{asymptomatic} = \frac{1}{15\%}$, $w_{symptomatic} = 1$ and $M = c \times w_{asymptomatic} + d \times w_{symptomatic}$.

We will report a bootstrap 95% confidence interval using stratified resampling. For each bootstrap iteration, we will resample with replacement within strata defined by clinical outcome (asymptomatic/symptomatic), preserving the observed proportion of infants with and without symptoms. The 95% confidence interval is calculated from 10,000 bootstrap replicates.

*Table 4. Incidence of neonatal infection*

| | Clinical outcome* | | |
|---|---|---|---|
| Baby result* | Negative | Positive | Total |
| Negative | a | b | a+b |
| Positive | c | d | c+d |
| Total | a+c | b+d | N |

* Order of test results is the same as Stata output when 0=negative, 1=positive

#### Cumulative incidence of neonatal infection by 4-6 weeks follow-up

We will estimate the cumulative incidence of any infection detected at any study visit. Cumulative incidence will be estimated using a Kaplan-Meier estimate with the following framework:

- **Time origin:** Date of birth.
- **Event time:** Date of first positivity.
- **Censoring:** Infants who complete all scheduled follow-up visits without infection will be censored at the date of their last negative test. Infants lost to follow-up will be censored at the date of their last attended visit.
- **Outcome:** Cumulative incidence will be calculated as 1 - probability of remaining infection-free at the end of follow-up, with 95% confidence interval. These probabilities will be reported at 28, 35 and 42 days after birth.

This framework will be used to identify the cumulative incidence of: *C. trachomatis* on eye swabs; *N. gonorrhoeae* on eye swabs*;* either *C. trachomatis* or *N. gonorrhoeae* on eye swabs; eye infection.

#### Diagnostic performance

We will assess the diagnostic accuracy of the GeneXpert CT/NG assay on eye and nasal swabs using Roche cobas CT/NG PCR as the reference standard, per pathogen and sample type. This analysis will be done on the **fully study population**, Paired swab results (Xpert and cobas PCR from the same baby, pathogen and sample type) from visits 1 and 2 will be included.

Separate tables will be made for each pathogen and sample type following the format in Table 5. Separate tables will also be made for babies with and without symptoms.

Diagnostic performance metrics will include overall concordance, sensitivity, specificity, positive predictive value (PPV) and negative predictive value (NPV). To account for repeated measurements within babies, each performance measure will be estimated using an intercept-only logistic regression model fitted to the relevant subset of paired test results (e.g., test positive among disease-positive samples for sensitivity). Estimates will be reported as marginal probabilities with 95% confidence intervals calculated using cluster-robust (sandwich) standard errors clustered on the baby's identification number.[4]

*Table 5. Diagnostic concordance*

| | cobas CT/NG PCR result* | | |
|---|---|---|---|
| Xpert CT/NG result* | Negative | Positive | Total |
| Negative | a | b | a+b |
| Positive | c | d | c+d |
| Total | a+c | b+d | N |

* Order of test results is the same as Stata output when 0=negative, 1=positive

### Statistical software

Analyses will be conducted with Stata 18.0 (or more recent) or R 4.4.2 (or more recent). AI-based coding assistants, such as GitHub Co-pilot, may be used to develop code.

WANTAIM

Text S2. WANTAIM Trial study team members and project staff

**WANTAIM Trial study team**

| | |
|---|---|
| Prof. Andrew Vallely | Professorial Research Fellow, Sexual and Reproductive Health, Papua New Guinea Institute of Medical Research, PO Box 60, Goroka, Eastern Highlands Province EHP 441 Papua New Guinea; and Associate Professor, Public Health Interventions Research Group, The Kirby Institute, University of New South Wales, Wallace Wurth Building, Sydney, NSW 2052, Australia<br>E: |
| Prof. William Pomat | Deputy Director, Science, and Head, Infection and Immunity Unit, Papua New Guinea Institute of Medical Research, PO Box 60, Goroka, Eastern Highlands Province EHP 441 Papua New Guinea<br>E: |
| Prof. Caroline Homer | Co-Program Director, Maternal and Child Health,<br>Burnet Institute, 85 Commercial Road, Melbourne, Victoria 3004 Australia<br>E: |
| Prof Rebecca Guy | Head, Surveillance Evaluation and Research Program, The Kirby Institute, University of New South Wales, Wallace Wurth Building, Sydney, NSW 2052, Australia<br>E: |
| Prof John Kaldor | Professor of Epidemiology and NHMRC Senior Principal Research Fellow,<br>Head, Public Health Interventions Research Group,<br>The Kirby Institute, University of New South Wales, Wallace Wurth Building, Sydney, NSW 2052, Australia<br>E: |
| A/Prof Stanley Luchters | Professor and Chair of the Department of Population Health, The Aga Khan University Sunny Plaza \| 4th Parklands Avenue off Wangapala Rd, Nairobi, Kenya.<br>E: |
| Prof Glen Mola (Trial Medical Expert) | Head of Reproductive Health, Obstetrics and Gynaecology, School of Medicine and Health Sciences, University of Papua New Guinea,<br>PO Box 5623 Boroko, NCD, Papua New Guinea<br>E: |
| Dr Grace Kariwiga (Trial Medical Expert ) | Consultant Obstetrician and Gynaecologist, Alotau Provincial Hospital, Milne Bay Provincial Health Authority, Locked Bag 402, Alotau, Milne Bay Province, Papua New Guinea<br>E: |
| Dr Lisa Vallely | Lecturer, International Maternal Health, Public Health Interventions Research Group, The Kirby Institute, University of New South Wales, Wallace Wurth Building, Sydney, NSW 2052, Australia<br>E: |
| Prof Virginia Wiseman (Trial Health Economist) | Health Economist, School of Public Health and Community Medicine, University of New South Wales, UNSW Sydney 2052, Australia<br>Health Economist/Senior Lecturer, Faculty of Public Health and Policy, Department of Global Health and Development, London School of Hygiene & Tropical Medicine, Keppel Street, London WC1E 7HT<br>E: |
| Dr Chris Morgan (Trial Health Systems Specialist) | Principal Fellow, Centre of International Health, Burnet Institute, Melbourne, Victoria, Australia<br>E: |

| | |
|---|---|
| A/Prof Handan Wand (Trial Statistician) | Biostatistics and Database Program, The Kirby Institute, University of New South Wales, Wallace Wurth Building, Sydney, NSW 2052, Australia<br>E: |
| Prof Stephen Rogerson | Department of Medicine (RMH/WH),<br>University of Melbourne, 4th Floor Clinical Science Building, Royal Parade, Parkville,<br>VIC 3050, Australia.<br>E: |
| Prof Sepehr Tabrizi | Department of Microbiology, Division of Laboratory Services, The Royal Women's Hospital / The Royal Children's Hospital, Melbourne Professor, Faculty of Medicine, Dentistry and Health, Department of Obstetrics and Gynaecology, University of Melbourne, Carlton 3053, Australia<br> |
| A/Prof David Whiley (Chair, Laboratory Sub-group) | NHMRC Career Development Fellow, The University of Queensland, UQ Centre for Clinical Research; Research Scientist, Microbiology, Pathology Queensland;<br>Building 71/918, Royal Brisbane Hospital, Herston Qld 4029, Australia<br> |
| Prof Suzanne Garland | Director of Microbiological Research and Head of Clinical Microbiology and Infectious Diseases Royal Women's Hospital,<br>Professor, Faculty of Medicine, Dentistry and Health, Department of Obstetrics and Gynaecology University of Melbourne<br>Senior Consultant, Microbiology, Royal Children's Hospital, Level 1, Building 404, Bio 21 Institute, 30 Flemington Road, Parkville, Vic. 3052 Australia<br> |
| Prof Nicola Low | Professor of Epidemiology and Public Health, Director of Research University of Bern, Finkenhubelweg 11, 3012 Bern, Switzerland<br>E: |
| Prof Rosanna Peeling | Professor and Chair of Diagnostics Research, Department of Clinical Research, London School of Hygiene & Tropical Medicine, Keppel Street London WC1E 7HT, UK<br>E: |
| Prof Peter Siba | Director, Research<br>Divine Word University<br>Madang 511 Papua New Guinea<br>E: |
| Dr Michaela Riddell (Trial Coordinator) | Clinical Trial Coordinator, Papua New Guinea Institute of Medical Research, PO Box 378, Madang 511, Papua New Guinea<br>E: |
| Dr Moses Laman | Deputy Director, Papua New Guinea Institute of Medical Research, PO Box 378, Madang 511, Papua New Guinea<br>E: |
| Dr John Bolnga | Consultant Obstetrician and Gynaecologist, Department of Obstetrics & Gynaecology, Modilon General Hospital, PO Box 1200, Madang, Papua New Guinea.<br>E: |
| Dr Leanne Robinson | Head, Vector Borne Disease Unit, Papua New Guinea Institute of Medical Research,<br>PO Box 378, Madang 511, Papua New Guinea<br>E: |

| | |
|---|---|
| Dr Jacob Morewaya | Head, Public Health Division, Milne Bay Provincial Health Authority, Locked Bag 402, Alotau, Milne Bay Province, Papua New Guinea<br>E: |
| Dr Steve Badman (Laboratory QC/QA Coordinator) | The Kirby Institute, UNSW Australia Wallace Wurth Building, Sydney, NSW 2052<br>E: |
| Dr Neha Batura | Research Associate (Economics)<br>Institute for Global Health, University College London<br>30 Guilford Street, London WC1N 1EH<br>E: |
| A/Prof Angela Kelly-Hanku | The Kirby Institute, University of New South Wales, Wallace Wurth Building, Sydney, NSW 2052, Australia<br>E: |
| Dr Pamela Toliman | Papua New Guinea Institute of Medical Research,<br>PO Box 60, Goroka, EHP 441, Papua New Guinea<br>E: |
| Mr Wilfred Peter | Communications and Public Relations Officer, Modilon General Hospital, PO Box 1200, Madang, Papua New Guinea.<br>E: |
| Dr Liz Peach | Research Program Manager - Healthy Mothers Healthy Babies<br>The Burnet Institute, Section 90 Allotment 1, Takubar<br>P.O.Box 1458, Kokopo, East New Britain Province, Papua New Guinea<br>E: |

**WANTAIM project staff in East New Britain and Madang Provinces and study sites**

**Trial administration**

| | |
|---|---|
| Kelvin Waukave | Financial Controller |
| Wilfred Peter | Communications Advisor (Madang Provincial Health Authority) |
| Jacob Morewaya | Public Health Advisor (Milne Bay Provincial Health Authority) |
| Peter Siba | Senior Technical Advisor |
| Elizabeth Peach | Research Manager (Burnet Institute) |
| Patricia Sengele | Administration/Finance Officer (Burnet Institute) |

**East New Britain Province staff**

| | |
|---|---|
| Lucy Au | Site Coordinator – East New Britain |
| Irene Pukai Gani | Site Coordinator – East New Britain |
| Leah Molok | Health Extension Officer/Research Nurse |
| Tessie Clip | Health Extension Officer /Research Nurse |
| Irene Daniels | Health Extension Officer /Ultrasound scan technician |
| Crystal Keiwaga | Health Extension Officer /Ultrasound scan technician |
| Daniel Hosea | Midwife/Research Nurse |
| Augustina Aiarak | Midwife/Research Nurse |
| Lorraine Mua | Midwife/Research Nurse |
| Valentine Russiat | Laboratory Officer/GeneXpert technician |
| Anna Davis | Research Nurse/GeneXpert technician |
| Biru Subey | Research Nurse/GeneXpert technician |
| Misilie Padik | Research Nurse |
| Muria Tangal | Research Nurse |
| Esleen Vovono | Research Nurse |

| | |
|---|---|
| Konsetta Malava | Research Nurse |
| Ellen Kavang | Research Nurse |
| Vicky Bayagau Wong | Research Nurse |
| Jermimah Garaen | Research Community Health Worker |
| Laniet Eddie | Community Health Worker |
| Johnslyne David | Community Health Worker |
| Diana Malip | Community Health Worker |
| Noel Amada | Driver |
| Elisha Jordan | Driver |
| Mosely Viringa | Driver |
| Cosmos Francis | Driver (Burnet Institute) |
| James Makap | Driver (Burnet Institute) |
| Daniel Amin | Laboratory Officer (Burnet Institute) |
| Ruth Fidelis | Senior Laboratory Officer (Burnet Institute) |
| Tony Rave | HEO/Community Liaison Officer |
| Rebecca Anian | Community Liaison Officer/Admin Officer |
| Romalus Tavui | Community Liaison Officer |
| John Kamit | Community Liaison Officer |
| Benedictor Mission | Community Liaison Officer |
| Charity Stanley | Community Liaison Officer |
| Grace Baining | Finance officer (Burnet Institute) |
| Everlyn Kavang | Human Resources officer (Burnet Institute) |

**Madang Province staff**

| | |
|---|---|
| Sharon Warel | Midwife/Research Nurse/USS Technician |
| Janeth Kulimbao | Community Health Worker /Ultrasound scan technician |
| Talitha Manie | Research Nurse / Ultrasound scan technician |
| Dupain Singirok | Senior Research Nurse |
| Carolyn Wokias Augusto | Senior Research Nurse/Community Liaison |
| Eunice Jally | Health Extension Officer/Research Nurse |
| Maggie Taupa | Research Nurse |
| Regina Enman | Research Nurse |
| Judith Demie | Research Nurse |
| Pamela Brea | Research Nurse |
| Aileen Jeffrey | Research Nurse |
| Jonathon Warel | Research Nurse |
| Cornelia Duba | Laboratory Officer/GeneXpert Technician |
| George Kuias | Research Nurse/GeneXpert Technician |
| Beromina Jano | Research Nurse/GeneXpert Technician |
| Michelyn John | Research Nurse/GeneXpert Technician |
| Georgina Sengum | Research Community Health Worker |
| Joyce Soalili | Research Community Health Worker |
| Francesca Buran | Research Community Health Worker |
| Cegatha Tawai | Research Community Health Worker |
| Milda Lasu | Research Community Health Worker |

| | |
|---|---|
| Tia Marie Badem | Community Liaison Officer |
| Theresa Tivud | Community Liaison Officer |
| Joseph Yamuna | Operations Manager |
| Mathilda Saki | Data Manager |
| Kelly Masil | Data Manager |
| Lennie Mal | Data Entry Officer |
| Yapi Kepea | Driver |
| Paul Romanus | Driver |
| Konnie Patrick | Driver/Community Liaison Officer |

**Study sites**

| | |
|---|---|
| Paparatava Health Centre | Antenatal Clinic, Labour/Postnatal, East New Britain (Catholic Health Services) |
| Warangoi Rural Hospital | Antenatal Clinic, Labour/Postnatal, East New Britain (Provincial Health Authority services) |
| Jomba Clinic | Antenatal Clinic, Madang (Provincial Health Authority services) |
| Mugil Health Centre | Antenatal Clinic, Labour/Postnatal, Madang (Catholic Health Services) |
| Yaguam Rural Hospital | Antenatal Clinic, Labour/Postnatal, Madang (Lutheran Health Services) |
| St Mary's Vunapope | Labour/Postnatal, East New Britain (Catholic Health Services) |
| Nonga General Hospital | Labour/Postnatal, East New Britain (Provincial Health Authority services) |
| Madang Provincial Hospital | Labour/Postnatal, Madang (Provincial Health Authority services) |
